# Excess risk and clusters of symptoms after COVID-19 in a large Norwegian cohort

**DOI:** 10.1101/2021.10.15.21265038

**Authors:** Ida Henriette Caspersen, Per Magnus, Lill Trogstad

**Author notes:** **Correspondence to:** Ida Henriette Caspersen, Centre for Fertility and Health, Norwegian Institute of Public Health, Postbox 222 Skøyen, N-0213 Oslo, Norway.

## Abstract

Physical, psychological and cognitive symptoms have been reported as post-acute sequelae for COVID-19 patients but are also common in the general, uninfected population. We aimed to calculate the excess risk and identify patterns of 22 symptoms up to 12 months after COVID-19 infection. We followed more than 70,000 participants in an ongoing cohort study, the Norwegian Mother, Father and Child Cohort Study (MoBa) during the COVID-19 pandemic. Infected and non-infected cohort participants registered presence of 22 different symptoms in March 2021. One year after the initial infection, 13 of 22 symptoms were associated with SARS-CoV-2 infection, based on relative risks between infected and uninfected subjects. For instance, 17.4% of SARS-CoV-2 infected cohort participants reported fatigue that persist 12 months after infection, compared to new occurrence of fatigue that had lasted less than 12 months in 3.8% of non-infected subjects (excess risk 13.6%). The adjusted relative risk for fatigue was 4.8 (95 % CI 3.5 to 6.7). Two main underlying factors explained 50% of the variance in the 13 symptoms. Brain fog, poor memory, dizziness, heart palpitations, and fatigue had high loadings on the first factor, while shortness-of breath and cough had high loadings on the second factor. Lack of taste and smell showed low to moderate correlation to other symptoms. Anxiety, depression and mood swings were not strongly related to COVID-19. Our results suggest that there are clusters of symptoms after COVID-19 due to different mechanisms and question whether it is meaningful to describe long COVID as one syndrome.

## INTRODUCTION

In light of the many million people infected by SARS-CoV-2, it is important to understand the long-term physical, psychological and cognitive consequences for infected subjects in a population perspective. How common are the symptoms that persist or occur after infection, how long will they last, and what do they consist of? Since reported symptoms are mostly of a general nature, apart from altered smell and taste, one must take account of the incidence of these complaints in the uninfected population. Recent reviews^1-3^ of post-acute COVID-19 syndrome or long COVID mostly refer to follow-up studies of patients treated in the specialised health services. These studies are important for detailed understanding of the multi-organ sequelae of COVID-19, but do not represent all infected subjects, do not take account of the incidence of symptoms in the general, uninfected population, and do not subtract symptoms that were present before infection. The reviews show that the design, sampling and outcome measures in follow-up studies are heterogeneous, making meta-analyses difficult^1-3^. There is a need for population-based, large cohort studies with long-term follow-up that registers new symptoms both in infected and non-infected subjects.

We have used the Norwegian Mother, Father and Child Cohort Study (MoBa)^4^, and compare the proportion of new symptoms for participants with and without COVID-19, calculating excess risks for each symptom. We also show the number of symptoms reported by each participant and examine the symptom profile for participants infected 1-6 or 11-12 months ago. We compare risks for men and women, and for subjects with and without severe disease.

There is no clear consensus of what constitutes the post-acute COVID-19 syndrome. Since there are symptoms from several organ systems, such as the nervous and respiratory systems, it is of interest to aggregate the long list of symptoms into clusters that can explain as much variation as possible. Using exploratory factor analysis, we have reduced the complexity of symptoms into two factors.

## METHODS

### Study design

The Norwegian Mother, Father and Child Cohort Study (MoBa) is a population-based pregnancy cohort study conducted by the Norwegian Institute of Public Health. Participants were recruited from all over Norway from 1999-2008.^4^ The women consented to participation in 41% of the pregnancies and the cohort now includes 95,000 mothers and 75,000 fathers. Parents and children have been followed with questionnaires and registry linkages with the aim to understand causes of disease. Using each participant’s unique national identification number, MoBa was linked to, the Medical Birth Registry of Norway (MBRN), the National Population Registry, the National Surveillance System for Communicable Diseases (MSIS)^5^ and to the Norwegian Immunisation Registry (SYSVAK).^6^ It is mandatory to report cases of COVID-19 to MSIS and vaccinations against SARS-CoV-2 to SYSVAK.

### Study population

Since March 2020, about 150,000 adult active cohort participants cohort participants have been invited to answer electronic questionnaires every 14 days with questions related to COVID-19. The response rates to the questionnaires distributed between March 2020 and March 2021 have been 50-80%. For the current study, eligible participants included all adult cohort members who were invited to answer a questionnaire in March 2021 (n=139,326) about current symptoms and, if present, the duration of such symptoms. The questions were posed to all cohort participants, regardless of previous COVID-19. An inclusion criterion was that participants had to be non-vaccinated against COVID-19 as vaccination was initiated in Norway in late December 2020. We therefore excluded participants who were registered with any vaccine dose against SARS-CoV-2 in SYSVAK if the vaccination date preceded or corresponded to the date when symptoms were reported (n=6844). We also excluded participants who received a COVID-19 diagnosis in February or March 2021 (n=486), to ensure that all COVID-19 cases in our study sample had been diagnosed at least 4 weeks earlier (Figure 1).

**Figure 1.**
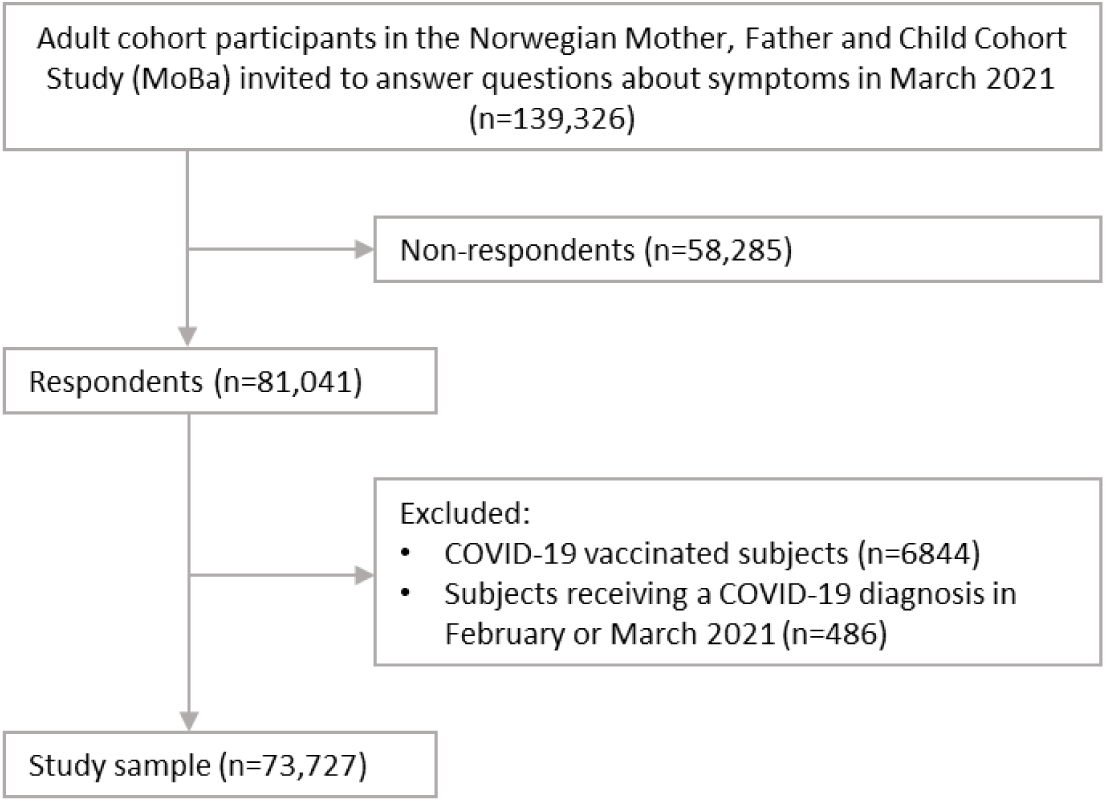
Flow chart of MoBa participants.

### Exposures

A COVID-19 diagnosis was obtained from registry data (MSIS) based on PCR confirmed SARS-CoV-2 infection^6^. We considered participants registered with a diagnosis before February 1^st^, 2021 as COVID-19 cases. Multiple registrations were not identified for any of the participants included in the study sample. Based on the two main waves of increased infection rates in Norway, we performed stratified analyses with i) a COVID-19 diagnosis in spring (Wave-1), including participants who were registered with a diagnosis in the period before May 1^st^ 2020 (first registration was March 6^th^), and ii) a COVID-19 diagnosis in autumn/winter (Wave-2), including those with a diagnosis registered between September 1^st^ 2020 and January 31^st^ 2021. The control groups were defined by those who had not received a COVID-19 diagnosis at all during the study period.

### Outcomes

A questionnaire was distributed to the participants’ mobile phones on March 2^nd^, 2021. The questionnaire included a list of 22 specific symptoms/signs or diseases (Supplementary Table 1). For simplicity, we refer to all listed symptoms/signs/diseases as “symptoms”. All participants were asked to check off if they had any of the listed symptoms. The symptoms were selected based on a list from Centres for Disease Control (CDC) in the US^7^. Participants reporting any symptom were also asked about the duration (<1 month, 1-3 months, 4-6 months, 7-12 months, 13-18 months, >18 months). Among COVID-19 cases, we considered symptoms to be novel if the reported duration was shorter than the time since acquiring a COVID-19 diagnosis.

### Covariates

Vaccination status was obtained from the Norwegian Immunisation Registry (SYSVAK)^12^. From the existing MoBa database, we included variables on the participants’ age (continuous, calculated from birth year), gender (defined by cohort member role as mother or father), and as proxy of socioeconomic status, educational level (less than high school, high school, college ≤4 years, more than 4 years college). Underlying chronic illness were self-reported in three questionnaires to cohort participants distributed in March and April 2020. The following diseases were reported: asthma or other lung disease, cancer, heart disease, hypertension, diabetes, other disease, or no disease. We grouped participants together if they reported at least one disease (including “other disease”) in any of the three questionnaires. From the ongoing data collection, we also included information from January 2021 on current smoking (no/yes; and if yes: occasional/daily) and body mass index (BMI, calculated from height and weight).

### Statistical analysis

We estimated associations between COVID-19 status and symptoms reported in March 2021 using log-binomial regression models with heteroscedasticity consistent (robust) standard errors. Associations were reported as excess risk (risk differences, RD) and relative risks (RR). RRs were reported with 95% confidence intervals (CI). We examined associations with unadjusted and adjusted regression models including age and chronic disease. We also examined associations after adjustment for education, BMI and smoking in addition to age and chronic illness. We performed analyses stratified by timing of infection (Wave 1 or 2) while excluding case subjects reporting symptom duration exceeding the time since infection. Controls were included based on recency of reported symptoms and excluded if they reported symptoms with duration >12 months when compared with cases in Wave-1 or >6 months when compared with cases in Wave-2. We also performed analyses stratified by self-reported severity of symptoms and gender, using any COVID-19 diagnosis before February 1^st^ 2021 as exposure.

We examined bivariate correlations between symptoms using tetrachoric correlations, assuming an underlying bivariate normal distribution of the symptoms. Significance of correlations were indicated by a correlation test inferring Pearson correlation with α=0.05. Symptom patterns among all COVID-19 cases were derived from exploratory factor analysis using the tetrachoric correlation matrix. To decide the number of factors, we used Horn’s Parallel Analysis for factor retention (Supplementary Figure 1). We examined both the two-factor and three-factor solutions, Supplementary Table 2. We also explored both direct oblimin (oblique, allowing correlated factors) and varimax (orthogonal) rotations. The two underlying factors identified were similar in the two rotation approaches (Table 5 and Supplementary Table 3). We omitted rare occurrences (myocarditis, kidney disease) from the factor analysis, as well as symptoms with no increased risk (based on RR) among COVID-19 cases 11-12 months after infection (sleep problems, depression, mood swings, joint pain, muscle pain, hair loss, and fever). Statistical analyses were done in R, version 4.1.0, using packages *tidyverse, sandwich, nFactor, polychor, cor*.*mtest, corrplot, psych*.

#### Missing data

The proportion of missing values in covariates was 11.5% for chronic illness and 3.7% for education level. We performed multiple imputation by chained equations with 20 imputations, using the R package *mice*. The dataset used for imputation included the following variables: chronic illness, education, age, BMI, smoking, gender, Covid-19 diagnosis, and long-term symptoms. Imputed values did not differ substantially from observed values. For instance, the proportion of chronic illness was 29.3% for imputed values and 29.4% in observed data. For education, the proportions in the four categories were as follows (imputed vs. observed): <High school 6.3% vs. 6.0%, high school 30.3% vs. 30.1%, college ≤4 years 37.9% vs. 37.7%, college >4 years 25.4% vs 26.2%. Tables 2-4 show results from analyses with imputed missing values in covariates. Complete case analyses for Wave-1 is presented in Supplementary Table 4.

## RESULTS

In total, 774 (1.0%) of the 73,727 included cohort participants were infected with SARS-CoV-2 in the study period. Figure 2 shows the number of infected MoBa participants per month. Of these, 170 were infected in March or April 2020 (defined as Wave-1 subjects), and 583 participants infected from September 2020 to January 2021 (Wave-2 subjects). All infected and non-infected participants constitute the study population (Table 1).

**Table 1.** Study sample characteristics. Data are numbers (%).

|  | No COVID-19 (n=72953)<sup>a</sup> | COVID-19 cases (n=774)<sup>b</sup> |
| --- | --- | --- |
| **Male** | 29658 (40.7) | 325 (42.0) |
| **Female** | 43295 (59.3) | 449 (58.0) |
| **Age (years)** |  |  |
| 25-34 | 489 (0.7) | 6 (0.8) |
| 35-39 | 5386 (7.4) | 58 (7.5) |
| 40-44 | 19525 (26.8) | 201 (26.0) |
| 45-49 | 27492 (37.7) | 292 (37.7) |
| 50-54 | 15245 (20.9) | 150 (19.4) |
| 55-59 | 3836 (5.3) | 53 (6.8) |
| 60-64 | 758 (1.0) | 10 (1.3) |
| 65+ | 222 (0.3) | 4 (0.5) |
| Missing | 0 (0) | 0 (0) |
| **Educational level** |  |  |
| < High school | 4230 (5.8) | 33 (4.3) |
| High school | 21144 (29.0) | 220 (28.4) |
| College ≤4 years | 26486 (36.3) | 298 (38.5) |
| College >4 years | 18401 (25.2) | 195 (25.2) |
| Missing | 2692 (3.7) | 28 (3.6) |
| **BMI** |  |  |
| <18.5 | 449 (0.6) | 2 (0.3) |
| 18.5-24.9 | 26973 (37.0) | 287 (37.1) |
| 25-29.9 | 23861 (32.7) | 263 (34.0) |
| 30-34.9 | 8219 (11.3) | 93 (12.0) |
| ≥35.0 | 2794 (3.8) | 31 (4.0) |
| Missing | 10657 (14.6) | 98 (12.7) |
| **Current smoking** |  |  |
| No | 58254 (79.9) | 645 (83.3) |
| Yes, occasional | 2000 (2.7) | 26 (3.4) |
| Yes, daily | 2861 (3.9) | 9 (1.2) |
| Missing | 9838 (13.5) | 94 (12.1) |
| **Chronic illness<sup>c</sup>** |  |  |
| No | 45564 (62.5) | 480 (62.0) |
| Yes | 18981 (26.0) | 197 (25.5) |
| Missing | 8408 (11.5) | 97 (12.5) |

<sup>a</sup> Participants not registered with a COVID-19 diagnosis before February 1<sup>st</sup> 2021. Participants with COVID-19 diagnosis in February or March 2021 were excluded from the study sample.
<sup>b</sup> Participants who were registered with a COVID-19 diagnosis in MSIS before February 1<sup>st</sup> 2021.
<sup>c</sup> Asthma or other lung disease, cancer, heart disease, hypertension, diabetes, other disease reported in March/April 2020.

**Figure 2.**
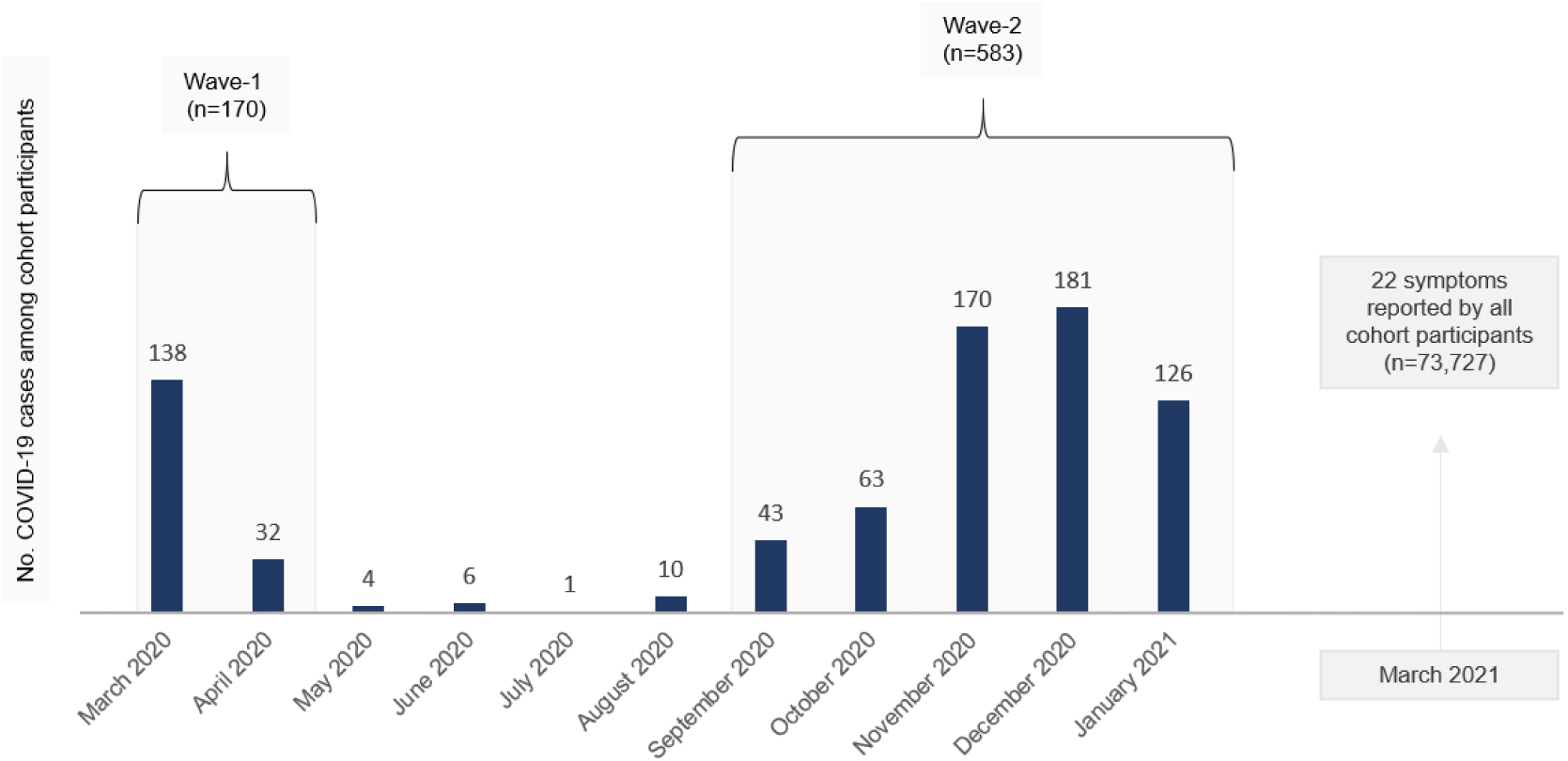
Incident cases of COVID-19 in the study sample, per month. The first registration of a COVID-19 diagnosis in our dataset was March 6^th^ 2020. We defined two waves of increased infection rates; March and April 2020 (Wave 1) and September 2020 to January 2021 (Wave 2). Symptoms were reported by all study participants in a questionnaire distributed March 2^nd^ 2021.

### Excess and relative risks after 11-12 months (Wave-1 subjects)

After 11-12 months, infected subjects had increased risk of 13 of the 22 symptoms when compared to uninfected subjects, with adjusted RRs ranging from 1.8 to 51.4 (Table 2). The symptom with highest excess risk (16.6%) 11-12 months after infection was altered smell or taste. The excess risk for this symptom is not much different from the absolute risk, since only 0.3% of the uninfected subjects reported this as a new symptom. This translates to a large adjusted relative risk (51.4, 95% CI: 36.0-73.5). Other symptoms with relatively high excess risks are poor memory (14.6%) and fatigue (13.6%). Psychological symptoms, such as anxiety, depression and mood swings have low excess risk, which is also the case for fever, muscle and joint pain. Reduced lung function (7.4%) and shortness-of breath (9.9%) have higher excess risk, and the adjusted relative risk of experiencing reduced lung function during the past year is as high as 24.9 (95% CI: 14.6-42.7), since very few (0.3%) uninfected subjects report this as a new symptom. The adjusted relative risk for chest pain was lower in complete case analysis (RR 4.2, 95% CI: 1.5-8.9) than in the imputed data analysis (RR 6.7, 95% CI: 3.6-12.7). Other symptoms showed no large changes in complete cases analysis. Relative risks showed no large changes after adjustment for education, BMI, and smoking in addition to age and prior chronic disease (Supplementary Table 4).

**Table 2.** Risks, excess risks (risk difference, RD) and relative risks (RR) for reporting current symptoms among cohort participants who acquired a COVID-19 diagnosis 11-12 months ago compared with controls with no COVID-19.

|  | n <sup>a, b</sup> | No COVID-19, n (%) with symptoms <sup>b</sup> | COVID-19 diagnosis 11-12 months ago, n (%) with symptoms <sup>b</sup> | RD | RR (95% CI), unadjusted | RR (95% CI), adjusted <sup>c</sup> |
| --- | --- | --- | --- | --- | --- | --- |
| **CARDIORESPIRATORY** |  |  |  |  |  |  |
| Chest pain | 73512 | 606 (0.8) | 9 (5.4) | 4.6 | 6.4 (3.4, 12.2) | 6.7 (3.6, 12.7) |
| Cough | 73231 | 1611 (2.2) | 8 (4.8) | 2.6 | 2.1 (1.1, 4.2) | 2.2 (1.1, 4.4) |
| Shortness of breath | 73257 | 963 (1.3) | 19 (11.2) | 9.9 | 8.4 (5.5, 12.9) | 8.7 (5.7, 13.3) |
| Heart palpitations | 72735 | 1478 (2.1) | 13 (7.7) | 5.6 | 3.8 (2.2, 6.4) | 3.9 (2.3, 6.6) |
| Myocarditis | 73718 | 5 (0) | 0 (0) | 0 | NA | NA |
| Reduced lung function | 73311 | 234 (0.3) | 13 (7.7) | 7.4 | 23.8 (13.9, 40.8) | 24.9 (14.6, 42.7) |
| **NEUROCOGNITIVE** |  |  |  |  |  |  |
| Anxiety | 72401 | 963 (1.3) | 6 (3.5) | 2.2 | 2.6 (1.2, 5.8) | 2.8 (1.3, 6) |
| Brain fog | 71516 | 2780 (3.9) | 20 (12) | 8.1 | 3.0 (2, 4.6) | 3.2 (2.1, 4.8) |
| Depression | 72364 | 2046 (2.9) | 7 (4.1) | 1.2 | 1.4 (0.7, 3.0) | 1.5 (0.7, 3.1) |
| Dizziness | 72652 | 2212 (3.1) | 10 (6) | 2.9 | 1.9 (1.1, 3.6) | 2.1 (1.1, 3.7) |
| Fatigue | 70956 | 2634 (3.8) | 29 (17.4) | 13.6 | 4.6 (3.3, 6.5) | 4.8 (3.5, 6.7) |
| Headache | 70742 | 4970 (7.1) | 20 (12) | 4.9 | 1.7 (1.1, 2.6) | 1.8 (1.2, 2.6) |
| Mood swings | 72571 | 3835 (5.3) | 11 (6.5) | 1.2 | 1.2 (0.7, 2.1) | 1.3 (0.7, 2.2) |
| Poor memory | 71578 | 2517 (3.6) | 30 (18.2) | 14.6 | 5.1 (3.7, 7.1) | 5.3 (3.8, 7.3) |
| Sleep problems | 69702 | 4436 (6.4) | 15 (9.3) | 2.9 | 1.4 (0.9, 2.3) | 1.5 (0.9, 2.4) |
| **JOINT AND MUSCLE** |  |  |  |  |  |  |
| Joint pain | 69583 | 1855 (2.7) | 7 (4.3) | 1.6 | 1.6 (0.8, 3.3) | 1.7 (0.8, 3.4) |
| Muscle pain | 69655 | 2623 (3.8) | 10 (6.1) | 2.3 | 1.6 (0.9, 2.9) | 1.7 (0.9, 3.0) |
| **OTHER** |  |  |  |  |  |  |
| Altered smell or taste | 73655 | 249 (0.3) | 28 (16.9) | 16.6 | 49.4 (34.5, 70.8) | 51.4 (36, 73.5) |
| Fever | 73580 | 344 (0.5) | 2 (1.2) | 0.7 | 2.5 (0.6, 9.9) | 2.6 (0.7, 10.5) |
| Hair loss | 73268 | 416 (0.6) | 1 (0.6) | 0 | 1.0 (0.1, 7.4) | 1.1 (0.2, 8.0) |
| Kidney disease | 73537 | 32 (0) | 0 (0) | 0 | NA | NA |
| Skin rash | 72784 | 1117 (1.6) | 6 (3.5) | 1.9 | 2.3 (1.0, 5.0) | 2.4 (1.1, 5.2) |

<sup>a</sup> Number of subjects included in regression model.
<sup>b</sup> Participants with symptom duration >12 months were excluded from both case and control groups.
<sup>c</sup> Adjusted for age and chronic illness.

### Excess and relative risks after 1-6 months (Wave-2 subjects)

The picture is much the same after 1-6 months (Table 3). The excess risk for altered smell or taste is 21.8%, while it is 11.7% for poor memory, 17.4% for fatigue and 14.2% for shortness-of-breath. Headache (excess risk 8.9%), dizziness (8.0%), muscle or joint pain (both 4.9%) appear to be relatively more common after 1-6 months compared to 11-12 months. Anxiety and depression have low excess risk also after 1-6 months.

**Table 3.** Risks, excess risks (risk difference, RD) and relative risks (RR) for reporting current symptoms among cohort participants who acquired a COVID-19 diagnosis 1-6 months ago compared with controls with no COVID-19.

|  | n <sup>a, b</sup> | No COVID-19, n (%) with symptoms | COVID-19 diagnosis 1-6 months ago, n (%) with symptoms <sup>b</sup> | RD | RR (95% CI), unadjusted | RR (95% CI), adjusted <sup>c</sup> |
| --- | --- | --- | --- | --- | --- | --- |
| **CARDIORESPIRATORY** |  |  |  |  |  |  |
| Chest pain | 73411 | 514 (0.7) | 31 (5.4) | 4.7 | 7.6 (5.3, 10.8) | 7.5 (5.2, 10.7) |
| Cough | 73103 | 1487 (2.1) | 41 (7.1) | 5 | 3.5 (2.6, 4.7) | 3.5 (2.6, 4.7) |
| Shortness of breath | 73052 | 777 (1.1) | 88 (15.3) | 14.2 | 14.2 (11.6, 17.4) | 13.9 (11.3, 17.1) |
| Heart palpitations | 72476 | 1229 (1.7) | 60 (10.5) | 8.8 | 6.1 (4.8, 7.8) | 6.1 (4.8, 7.8) |
| Myocarditis | 73716 | 3 (0) | 1 (0.2) | 0.2 | 41.7 (4.3, 400.4) | 42.3 (4.5, 396) |
| Reduced lung function | 73239 | 174 (0.2) | 32 (5.5) | 5.3 | 23 (15.9, 33.2) | 22.5 (15.5, 32.5) |
| **NEUROCOGNITIVE** |  |  |  |  |  |  |
| Anxiety | 72119 | 690 (1) | 14 (2.5) | 1.5 | 2.6 (1.5, 4.3) | 2.6 (1.5, 4.3) |
| Brain fog | 70942 | 2227 (3.2) | 84 (14.8) | 11.6 | 4.7 (3.8, 5.7) | 4.7 (3.8, 5.7) |
| Depression | 71911 | 1602 (2.3) | 22 (3.9) | 1.6 | 1.7 (1.1, 2.6) | 1.7 (1.1, 2.6) |
| Dizziness | 72414 | 1984 (2.8) | 62 (10.8) | 8.0 | 3.9 (3.1, 5) | 3.9 (3.1, 4.9) |
| Fatigue | 70514 | 2222 (3.2) | 116 (20.6) | 17.4 | 6.5 (5.5, 7.7) | 6.4 (5.4, 7.5) |
| Headache | 70367 | 4607 (6.6) | 88 (15.5) | 8.9 | 2.3 (1.9, 2.8) | 2.3 (1.9, 2.8) |
| Mood swings | 71830 | 3105 (4.4) | 38 (6.6) | 2.2 | 1.5 (1.1, 2.1) | 1.5 (1.1, 2.1) |
| Poor memory | 70831 | 1798 (2.6) | 81 (14.3) | 11.7 | 5.6 (4.5, 6.8) | 5.6 (4.5, 6.8) |
| Sleep problems | 68819 | 3569 (5.2) | 63 (11.4) | 6.2 | 2.2 (1.7, 2.8) | 2.2 (1.7, 2.8) |
| **JOINT AND MUSCLE** |  |  |  |  |  |  |
| Joint pain | 69205 | 1482 (2.2) | 39 (7.1) | 4.9 | 3.3 (2.4, 4.4) | 3.2 (2.4, 4.4) |
| Muscle pain | 69249 | 2225 (3.2) | 45 (8.1) | 4.9 | 2.5 (1.9, 3.3) | 2.5 (1.9, 3.3) |
| **OTHER** |  |  |  |  |  |  |
| Altered smell or taste | 73555 | 177 (0.2) | 128 (22) | 21.8 | 90.5 (73.2, 111.9) | 89.9 (72.7, 111.1) |
| Fever | 73539 | 304 (0.4) | 8 (1.4) | 1 | 3.3 (1.6, 6.6) | 3.3 (1.6, 6.6) |
| Hair loss | 73143 | 294 (0.4) | 16 (2.8) | 2.4 | 6.8 (4.1, 11.2) | 6.8 (4.2, 11.2) |
| Kidney disease | 73531 | 26 (0) | NA (NA) | NA | 0 (0, 0) | 0 (0, 0) |
| Skin rash | 72622 | 961 (1.3) | 22 (3.8) | 2.5 | 2.8 (1.9, 4.3) | 2.8 (1.9, 4.3) |

<sup>a</sup> Number of subjects included in regression model.
<sup>b</sup> Participants with symptom duration >6 months were excluded from both case and control groups.
<sup>c</sup> Adjusted for age and chronic illness.

### Risks according to mild or severe infection

The participants were asked whether they had been almost not ill, moderately ill, very ill or hospitalized during the initial infection with SARS-CoV-2. In Table 4 we include all subjects with COVID-19 (wave 1 and 2 and the few subjects infected between the waves) and compare subjects who reported almost no illness (mild illness) with subjects reporting more severe illness (moderately, very ill, or hospitalized). In general, the prevalence of symptoms is about twice as high for subjects with severe infection. For brain fog, the prevalence is 18.1% for severely ill and 9.2% for mildly ill subjects. For shortness-of-breath the proportions are 19.5% and 6.9%, while the figures for altered smell or taste are 23.5% and 16.0%, respectively.

**Table 4.** Prevalence, excess risks (risk difference, RD) and relative risks (RR) for reporting current symptoms among cohort participants who acquired a COVID-19 diagnosis, comparing mild and moderate/severe cases.

|  | n | Mild COVID-19, n<br>(%) with symptoms | Severe COVID-19, n<br>(%) with symptoms | RD | RR (95% CI),<br>unadjusted | RR (95% CI),<br>adjusted |
| --- | --- | --- | --- | --- | --- | --- |
| CARDIORESPIRATORY |  |  |  |  |  |  |
| Chest pain | 767 | 5 (1.5) | 35 (8.4) | 6.9 | 5.6 (2.2, 14.1) | 5.4 (2.1, 13.7) |
| Cough | 764 | 11 (3.3) | 38 (9.1) | 5.8 | 2.7 (1.4, 5.3) | 2.9 (1.5, 5.5) |
| Shortness of breath | 768 | 23 (6.9) | 82 (19.5) | 12.6 | 2.8 (1.8, 4.4) | 2.9 (1.9, 4.5) |
| Heart palpitations | 762 | 16 (4.8) | 58 (13.9) | 9.1 | 2.9 (1.7, 4.9) | 2.9 (1.7, 5.0) |
| Myocarditis | 774 | 0 (0) | 1 (0.2) | 0.2 | NA | NA |
| Reduced lung function | 770 | 9 (2.7) | 34 (8) | 5.3 | 2.9 (1.4, 6.1) | 2.8 (1.3, 5.8) |
| NEUROCOGNITIVE |  |  |  |  |  |  |
| Anxiety | 757 | 7 (2.1) | 14 (3.4) | 1.3 | 1.6 (0.6, 3.9) | 1.5 (0.6, 3.6) |
| Brain fog | 753 | 30 (9.2) | 74 (18.1) | 8.9 | 2.0 (1.3, 2.9) | 1.9 (1.3, 2.9) |
| Depression | 759 | 10 (3.0) | 21 (5.1) | 2.1 | 1.7 (0.8, 3.5) | 1.7 (0.8, 3.6) |
| Dizziness | 760 | 18 (5.5) | 51 (12.3) | 6.8 | 2.2 (1.3, 3.8) | 2.3 (1.4, 3.8) |
| Fatigue | 749 | 33 (10.2) | 111 (27) | 16.8 | 2.6 (1.8, 3.8) | 2.6 (1.8, 3.7) |
| Headache | 754 | 28 (8.6) | 80 (19.4) | 10.8 | 2.2 (1.5, 3.4) | 2.3 (1.5, 3.4) |
| Mood swings | 765 | 14 (4.2) | 35 (8.4) | 4.2 | 2.0 (1.1, 3.6) | 1.8 (1.0, 3.4) |
| Poor memory | 752 | 23 (7.1) | 89 (21.7) | 14.6 | 3.1 (2.0, 4.7) | 3.0 (1.9, 4.6) |
| Sleep problems | 731 | 27 (8.5) | 50 (12.6) | 4.1 | 1.5 (0.9, 2.3) | 1.5 (0.9, 2.3) |
| JOINT AND MUSCLE |  |  |  |  |  |  |
| Joint pain | 735 | 13 (4) | 34 (8.6) | 4.6 | 2.1 (1.1, 4.0) | 2.2 (1.2, 4.2) |
| Muscle pain | 738 | 9 (2.8) | 47 (11.7) | 8.9 | 4.2 (2.1, 8.4) | 4.3 (2.1, 8.8) |
| OTHER |  |  |  |  |  |  |
| Altered smell or taste | 769 | 53 (16.0) | 99 (23.5) | 7.5 | 1.5 (1.1, 2.0) | 1.4 (1.1, 1.9) |
| Fever | 773 | 1 (0.3) | 9 (2.1) | 1.8 | 7.1 (0.9, 55.8) | 7.5 (0.9, 61.1) |
| Hair loss | 766 | 2 (0.6) | 16 (3.8) | 3.2 | 6.3 (1.5, 27.4) | 6.8 (1.6, 29.1) |
| Kidney disease | 773 | 0 (0) | 0 (0) | 0 | NA | NA |
| Skin rash | 771 | 4 (1.2) | 22 (5.2) | 4 | 4.3 (1.5, 12.4) | 4.3 (1.5, 12.2) |

<sup>a</sup> Number of subjects in analysis, excluding those with symptoms lasting for more than 12 months.
<sup>b</sup> Adjusted for age, chronic illness, and timing of infection (Wave-1, summer, Wave-2).

### Risks according to gender

Women who have been infected by SARS-CoV-2 report higher prevalence of heart palpitations than infected men (14.1% vs 4.9%) (Supplementary Table 5). The relative risk is 2.9 (95% CI: 1.6, 5.5) adjusted for age, prior chronic disease, and severity of infection. They also report higher prevalence of brain fog, fatigue, headache, dizziness, poor memory and altered smell or taste.

### Number of symptoms

The proportion of Wave-1 subjects who have no symptoms after 11-12 months is 44%, while it is 38% for Wave-2 subjects after 1-6 months. Among uninfected subjects, the proportion without new symptoms during the last 12 months is 79% (Supplementary Table 6). Reporting more than 4 different symptoms was uncommon after both wave 1 and 2.

### Correlation between symptoms

Figure 3 shows bivariate correlations between the symptoms that are significantly associated to sequelae after COVID-19, for Wave-1 (left part) and Wave-2 subjects (right part). Altered smell or taste was weakly correlated with other symptoms. All other symptoms display one or more significant correlations with other symptoms, and the two figures suggest a similar pattern of symptoms for both waves. Using the correlation matrix from both Wave-1 and Wave-2 together (Supplementary Figure 1 and Supplementary Table 7), we found that two underlying, latent factors explained 33% and 17% (in total 50%) of the variance in symptoms (Table 5). The correlation between the two factors was 58%. The symptoms that load highest on the first factor are brain fog, poor memory, dizziness, heart palpitations, and fatigue, while shortness-of breath and cough load highest to the second factor.

**Figure 3.**
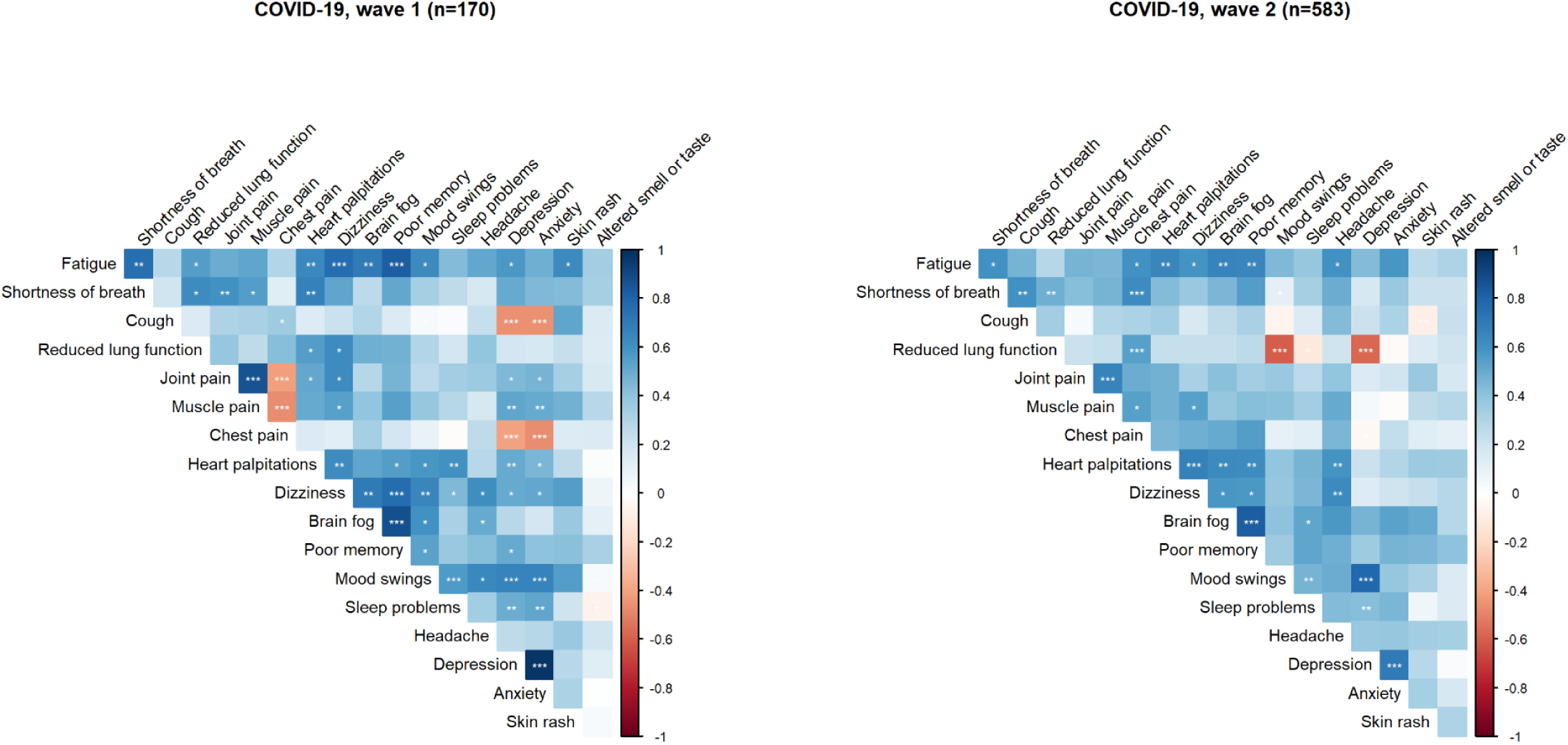
Bivariate tetrachoric correlations between symptoms reported in March 2021 among COVID-19 cases in wave 1 (11-12 months prior to reporting symptoms) and wave 2 (1-6 months prior to reporting symptoms). Rare occurrences (kidney disease, myocarditis, fever, hair loss) were omitted. The strength of correlation coefficients are indicated by the colour panel (right). Intensity of red colours indicate increasing negative correlation coefficients, while intensity of blue colours indicate increasing positive correlation coefficients. Correlation coefficients are found in Supplementary Table 7. Asterisks indicating significant correlations (*** for p<.001; ** for p<.01; * for p<.05).

**Table 5.** Loadings of an exploratory factor analysis of post-acute symptoms among COVID-19 cases (n=774). The analysis includes symptoms with increased risk among COVID-19 cases 11-12 months after diagnosis. The table shows standardized loadings from the pattern matrix using an oblimin rotation, allowing correlation between factors. The two factors explained 33% and 17% (in total 50%) of the variance in symptoms. ^a^

|  | **Factor1** | **Factor2** |
| --- | --- | --- |
| Brain fog | 0.95 | -0.11 |
| Poor memory | 0.82 | 0.10 |
| Dizziness | 0.81 | -0.11 |
| Heart palpitations | 0.74 | 0.05 |
| Fatigue | 0.62 | 0.30 |
| Headache | 0.57 | 0.18 |
| Skin rash | 0.45 | 0.02 |
| Anxiety | 0.41 | 0.15 |
| Altered smell or taste | 0.34 | 0.07 |
| Chest pain | 0.21 | 0.45 |
| Shortness of breath | 0.02 | 0.96 |
| Reduced lung function | 0.02 | 0.58 |
| Cough | -0.05 | 0.60 |

<sup>a</sup> We omitted rare occurrences (myocarditis, kidney disease) and symptoms with no increased risk (based on adjusted RR) among COVID-19 cases 11-12 months after infection (depression, mood swings, sleep problems, joint pain, muscle pain, fever, and hair loss).

## DISCUSSION

We have calculated the prevalence of symptoms after waves 1 and 2 of SARS-CoV-2 infection in Norway among subjects who have and have not been infected. This allows for measures of association between infection and symptoms, using both risk differences and relative risks. We find that some symptoms were clearly associated to infection with SARS-CoV-2. These symptoms cluster into sets, such as a neurocognitive set (e.g. brain fog, dizziness and poor memory) and a cardiorespiratory set (e.g. shortness-of-breath and cough). Altered smell or taste represent a frequent symptom that is more common in women and more common after severe infection but seems to have relatively low correlation to other symptoms. The findings support the view that long COVID may be more than one syndrome^8,9^. The excess risks for infected subjects were largest for altered smell or taste, poor memory, fatigue and shortness-of-breath after 11-12 months. The same symptoms had high excess risks after 1-6 months. It is interesting that muscle and joint pain have increased relative risk after 1-6 months, but not after 11-12 months.

An important finding is that there is low excess risk for anxiety and depression. Post-illness studies of patients with severe coronavirus infection have found a relatively high prevalence of depression^10^. We found that the risk of depression is higher for subjects who experienced a more severe infection initially compared to mildly infected subjects (adjusted RR 1.7, Table 4). The results suggest that anxiety and depression might be more a consequence of the severity of initial disease and the trauma of hospitalisation, rather than a direct consequence of the viral infection itself.

A unique feature of infection with SARS-CoV-2 is the change in smell and taste^11^. For many patients, these changes disappear shortly after the acute infection, but for a subgroup they remain. We find that 16.6% of Wave-1 subjects report altered smell or taste one year later. This is in accordance with proportions found across other studies^1-3^. The mechanisms behind this symptom is unknown. It is interesting that the correlations between altered smell or taste and cognitive symptoms in our study are relatively low, compatible with a suggestion that the main mechanism may be a local infection in olfactory epithelial cells^12^, rather than an intracerebral affection.

This study has limitations. If we compare the prevalence of symptoms in Wave-1 and Wave-2 subjects, and make inferences about the duration of symptoms, there are important assumptions. One is that the type of coronavirus may have changed over time, so that the infection, at least theoretically, may have different long-term consequences. The other factor is that testing opportunities were more restricted during the spring of 2020 compared to later months, possibly inducing differential selection for Wave-1 and Wave-2 subjects. These limitations will be overcome as we continue follow-up in MoBa, since viral sequencing has become prevalent, and the same individuals will be followed.

The symptoms in this study are self-reported in questionnaires, which has both advantages and draw-backs. The advantage is that one can ask a large, representative sample simple questions in a design that measures the prevalence, duration and correlational structure of symptoms, as well as the degree of association to prior infection. This type of data provides a supplement to what can be learned from linking population samples to health care registrations, as is done, for instance, in the follow-up of veterans in the US^13^. The draw-back is the lack of detail for each participant. There is a possibility for misclassification for signs or diseases that are not easily recognized by the study participants, like myocarditis and kidney disease, as we did not have objective measurements to complement self-reported data. In future research one can link to patient registries and select random subgroups from population-based cohorts for in-depth clinical investigation, in parallel with clinical follow-up of the more severely affected patients. This will give a fuller picture of the clinical spectrum of long COVID. A next step for large cohorts will be to understand why some infected subjects develop long COVID and others do not, by performing nested case-control studies that utilise biomaterials, including whole-genome genotyping.

In conclusion, the present design sheds light on the causal link between infection with SARS-CoV-2 and long-term symptoms and suggest distinct clusters of symptoms due to different effects of the virus. The lack of correlations between symptoms question whether the diverse manifestations after infection with SARS-CoV-2 can be classified as one syndrome.

### Ethical approvement

The establishment of MoBa and initial data collection was based on a license from the Norwegian Data Protection Agency and approval from The Regional Committees for Medical and Health Research Ethics. The MoBa cohort is now based on regulations related to the Norwegian Health Registry Act. The current sub-study was approved by The Regional Committee for Medical and Health Research Ethics, South East Norway C, no. 127708.

## Data Availability

Data from the cohort is available for analysis after approval from a Norwegian ethics committee and application to the Norwegian Institute of Public Health.

## Acknowledgements

The Norwegian Mother, Father and Child Cohort Study is supported by the Norwegian Ministry of Health and Care Services and the Ministry of Education and Research. We are grateful to all the participating families in Norway who take part in this on-going cohort study.

## Competing interests

The authors declare no competing interests.

## Funding

This work was funded by the Norwegian Research Council’s Centres of Excellence Funding Scheme (no. 262700) and by the Norwegian Institute of Public Health (NIPH).

## SUPPLEMENTARY MATERIAL

**Supplementary Table 1.**
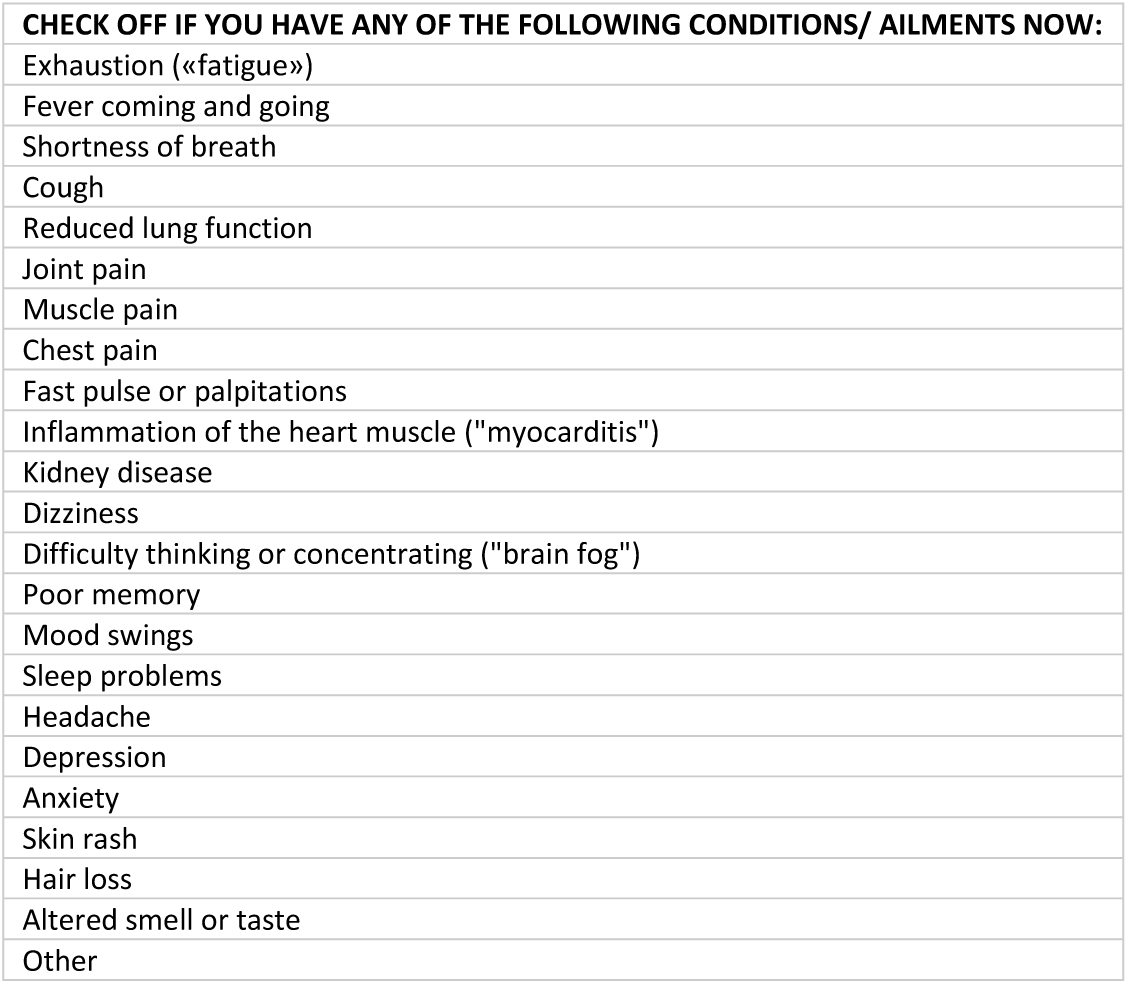
List of symptoms reported by all adult cohort participants independent of COVID-19 infection status.

**Supplementary Table 2.**
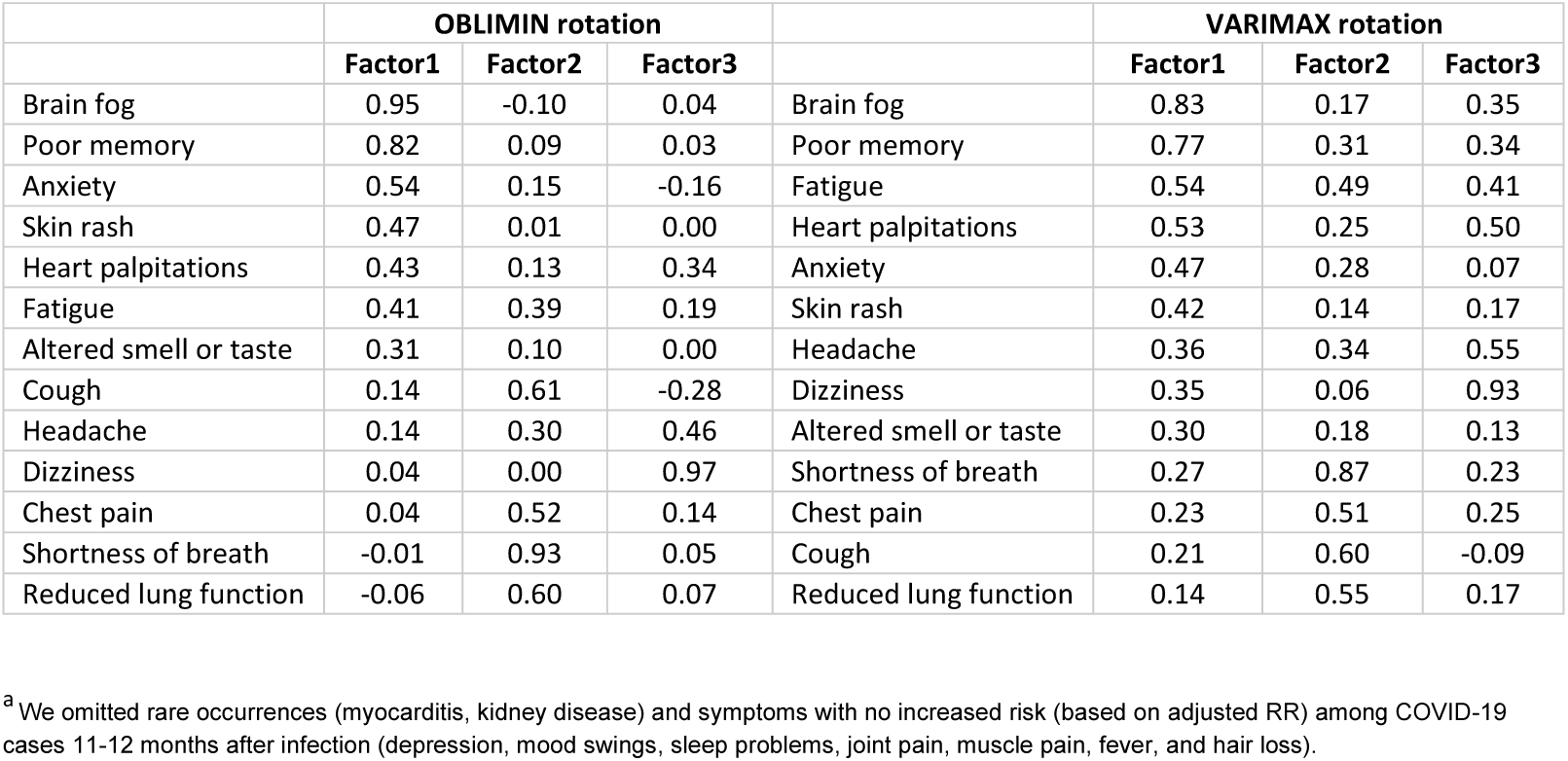
Loadings of three extracted factors of an exploratory factor analysis of post-acute symptoms among COVID-19 cases (n=774). The first three factors with oblimin rotation explained 55% of the total variation. and The analysis includes symptoms with increased risk among COVID-19 cases 11-12 months after diagnosis.^a^

|  | OBLIMIN rotation |  |  |  | VARIMAX rotation |  |  |
| --- | --- | --- | --- | --- | --- | --- | --- |
|  | Factor1 | Factor2 | Factor3 |  | Factor1 | Factor2 | Factor3 |
| Brain fog | 0.95 | -0.10 | 0.04 | Brain fog | 0.83 | 0.17 | 0.35 |
| Poor memory | 0.82 | 0.09 | 0.03 | Poor memory | 0.77 | 0.31 | 0.34 |
| Anxiety | 0.54 | 0.15 | -0.16 | Fatigue | 0.54 | 0.49 | 0.41 |
| Skin rash | 0.47 | 0.01 | 0.00 | Heart palpitations | 0.53 | 0.25 | 0.50 |
| Heart palpitations | 0.43 | 0.13 | 0.34 | Anxiety | 0.47 | 0.28 | 0.07 |
| Fatigue | 0.41 | 0.39 | 0.19 | Skin rash | 0.42 | 0.14 | 0.17 |
| Altered smell or taste | 0.31 | 0.10 | 0.00 | Headache | 0.36 | 0.34 | 0.55 |
| Cough | 0.14 | 0.61 | -0.28 | Dizziness | 0.35 | 0.06 | 0.93 |
| Headache | 0.14 | 0.30 | 0.46 | Altered smell or taste | 0.30 | 0.18 | 0.13 |
| Dizziness | 0.04 | 0.00 | 0.97 | Shortness of breath | 0.27 | 0.87 | 0.23 |
| Chest pain | 0.04 | 0.52 | 0.14 | Chest pain | 0.23 | 0.51 | 0.25 |
| Shortness of breath | -0.01 | 0.93 | 0.05 | Cough | 0.21 | 0.60 | -0.09 |
| Reduced lung function | -0.06 | 0.60 | 0.07 | Reduced lung function | 0.14 | 0.55 | 0.17 |

<sup>a</sup> We omitted rare occurrences (myocarditis, kidney disease) and symptoms with no increased risk (based on adjusted RR) among COVID-19 cases 11-12 months after infection (depression, mood swings, sleep problems, joint pain, muscle pain, fever, and hair loss).

**Supplementary Table 3.** Loadings of two extracted factors from an exploratory factor analysis of post-acute symptoms among COVID-19 cases (n=774), using varimax rotation. The analysis includes symptoms with increased risk among COVID-19 cases 11-12 months after diagnosis. The two factors explained 31% and 19% (in total 50%) of the variance in symptoms. ^a^

|  | VARIMAX rotation |  |
| --- | --- | --- |
|  | Factor1 | Factor2 |
| Brain fog | 0.87 | 0.19 |
| Poor memory | 0.80 | 0.34 |
| Dizziness | 0.74 | 0.15 |
| Heart palpitations | 0.72 | 0.28 |
| Fatigue | 0.68 | 0.48 |
| Headache | 0.60 | 0.35 |
| Anxiety | 0.44 | 0.27 |
| Skin rash | 0.44 | 0.16 |
| Altered smell or taste | 0.34 | 0.17 |
| Chest pain | 0.33 | 0.49 |
| Shortness of breath | 0.31 | 0.93 |
| Reduced lung function | 0.19 | 0.55 |
| Cough | 0.14 | 0.56 |

<sup>a</sup> We omitted rare occurrences (myocarditis, kidney disease) and symptoms with no increased risk (based on adjusted RR) among COVID-19 cases 11-12 months after infection (depression, mood swings, sleep problems, joint pain, muscle pain, fever, and hair loss).

**Supplementary Table 4.** Risks, excess risks (risk difference, RD) and adjusted relative risks (RR) for reporting current symptoms among cohort participants who acquired a COVID-19 diagnosis 11-12 months ago and controls with no COVID-19.

|  | Complete case data |  | Imputed data |  |
| --- | --- | --- | --- | --- |
|  | n <sup>a</sup> | RR (95% CI),<br>minimal<br>adjustment <sup>b</sup> | n <sup>a</sup> | RR (95% CI),<br>full adjustment <sup>c</sup> |
| CIRCULATION/RESPIRATION |  |  |  |  |
| Chest pain | 63009 | 4.2 (1.5, 8.9) | 73512 | 7.0 (3.7, 13.3) |
| Cough | 62764 | 2.5 (1.2, 4.6) | 73231 | 2.3 (1.2, 4.6) |
| Dyspnea | 62786 | 8.9 (5.4, 13.3) | 73257 | 9.4 (6.1, 14.3) |
| Heart palpitations | 62322 | 4.5 (2.6, 7.2) | 72735 | 4.0 (2.4, 6.8) |
| Myocarditis | 63183 | NA | 73718 | 0 (0, 0) |
| Reduced lung function | 62823 | 20.0 (9.7, 35.5) | 73311 | 25.9 (15, 44.8) |
| BRAIN |  |  |  |  |
| Anxiety | 62065 | 3.2 (1.3, 6.3) | 72401 | 2.9 (1.3, 6.2) |
| Brain fog | 61262 | 3.1 (1.9, 4.6) | 71516 | 3.2 (2.2, 4.8) |
| Depression | 62042 | 1.7 (0.7, 3.3) | 72364 | 1.6 (0.8, 3.2) |
| Dizziness | 62237 | 1.6 (0.7, 3.1) | 72652 | 2.1 (1.1, 3.8) |
| Fatigue | 60765 | 5.1 (3.5, 6.9) | 70956 | 4.9 (3.6, 6.8) |
| Headache | 60583 | 1.7 (1.0, 2.5) | 70742 | 1.8 (1.2, 2.7) |
| Mood swings | 62186 | 1.3 (0.7, 2.2) | 72571 | 1.3 (0.7, 2.2) |
| Poor memory | 61318 | 5.2 (3.6, 7.2) | 71578 | 5.4 (3.9, 7.5) |
| Sleep problems | 59698 | 1.5 (0.9, 2.4) | 69702 | 1.5 (0.9, 2.4) |
| JOINT AND MUSCLE |  |  |  |  |
| Joint pain | 59584 | 1.9 (0.8, 3.6) | 69583 | 1.7 (0.8, 3.6) |
| Muscle pain | 59646 | 1.9 (1.0, 3.2) | 69655 | 1.7 (0.9, 3.1) |
| OTHER |  |  |  |  |
| Altered smell or taste | 73052 | 52.3 (34.9, 74.1) | 73655 | 52.4 (36.6, 74.9) |
| Fever | 63065 | 1.5 (0.1, 6.6) | 73580 | 2.8 (0.7, 11.3) |
| Hair loss | 72667 | NA | 73268 | 1.1 (0.2, 8.1) |
| Kidney disease | 63021 | NA | 73537 | NA |
| Skin rash | 62377 | 2.2 (0.8, 4.7) | 72784 | 2.4 (1.1, 5.2) |

<sup>a</sup> Total number included in regression models.
<sup>b</sup> Adjusted for age and chronic illness.
<sup>c</sup> Adjusted for age, chronic illness, BMI, education, and smoking.

**Supplementary Table 5.**
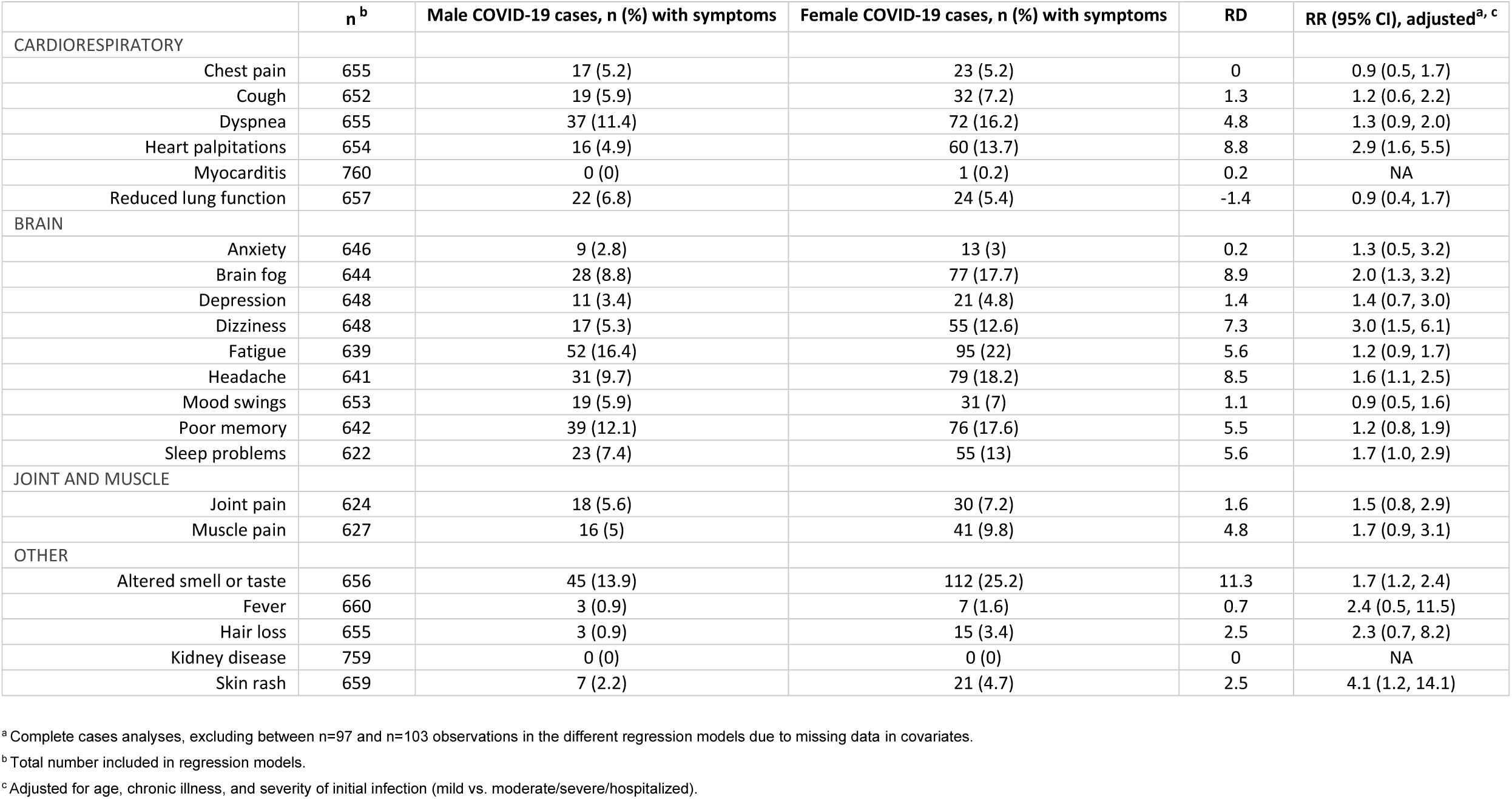
Risks, excess risks (risk difference, RD) and relative risks (RR) for reporting current symptoms for male versus female COVID-19 cases. Complete case analyses^a^.

|  | n<sup>b</sup> | Male COVID-19 cases, n (%) with symptoms | Female COVID-19 cases, n (%) with symptoms | RD | RR (95% CI), adjusted<sup>a, c</sup> |
| --- | --- | --- | --- | --- | --- |
| CARDIORESPIRATORY |  |  |  |  |  |
| Chest pain | 655 | 17 (5.2) | 23 (5.2) | 0 | 0.9 (0.5, 1.7) |
| Cough | 652 | 19 (5.9) | 32 (7.2) | 1.3 | 1.2 (0.6, 2.2) |
| Dyspnea | 655 | 37 (11.4) | 72 (16.2) | 4.8 | 1.3 (0.9, 2.0) |
| Heart palpitations | 654 | 16 (4.9) | 60 (13.7) | 8.8 | 2.9 (1.6, 5.5) |
| Myocarditis | 760 | 0 (0) | 1 (0.2) | 0.2 | NA |
| Reduced lung function | 657 | 22 (6.8) | 24 (5.4) | -1.4 | 0.9 (0.4, 1.7) |
| BRAIN |  |  |  |  |  |
| Anxiety | 646 | 9 (2.8) | 13 (3) | 0.2 | 1.3 (0.5, 3.2) |
| Brain fog | 644 | 28 (8.8) | 77 (17.7) | 8.9 | 2.0 (1.3, 3.2) |
| Depression | 648 | 11 (3.4) | 21 (4.8) | 1.4 | 1.4 (0.7, 3.0) |
| Dizziness | 648 | 17 (5.3) | 55 (12.6) | 7.3 | 3.0 (1.5, 6.1) |
| Fatigue | 639 | 52 (16.4) | 95 (22) | 5.6 | 1.2 (0.9, 1.7) |
| Headache | 641 | 31 (9.7) | 79 (18.2) | 8.5 | 1.6 (1.1, 2.5) |
| Mood swings | 653 | 19 (5.9) | 31 (7) | 1.1 | 0.9 (0.5, 1.6) |
| Poor memory | 642 | 39 (12.1) | 76 (17.6) | 5.5 | 1.2 (0.8, 1.9) |
| Sleep problems | 622 | 23 (7.4) | 55 (13) | 5.6 | 1.7 (1.0, 2.9) |
| JOINT AND MUSCLE |  |  |  |  |  |
| Joint pain | 624 | 18 (5.6) | 30 (7.2) | 1.6 | 1.5 (0.8, 2.9) |
| Muscle pain | 627 | 16 (5) | 41 (9.8) | 4.8 | 1.7 (0.9, 3.1) |
| OTHER |  |  |  |  |  |
| Altered smell or taste | 656 | 45 (13.9) | 112 (25.2) | 11.3 | 1.7 (1.2, 2.4) |
| Fever | 660 | 3 (0.9) | 7 (1.6) | 0.7 | 2.4 (0.5, 11.5) |
| Hair loss | 655 | 3 (0.9) | 15 (3.4) | 2.5 | 2.3 (0.7, 8.2) |
| Kidney disease | 759 | 0 (0) | 0 (0) | 0 | NA |
| Skin rash | 659 | 7 (2.2) | 21 (4.7) | 2.5 | 4.1 (1.2, 14.1) |

<sup>a</sup> Complete cases analyses, excluding between n=97 and n=103 observations in the different regression models due to missing data in covariates.
<sup>b</sup> Total number included in regression models.
<sup>c</sup> Adjusted for age, chronic illness, and severity of initial infection (mild vs. moderate/severe/hospitalized).

**Supplementary Table 6.**
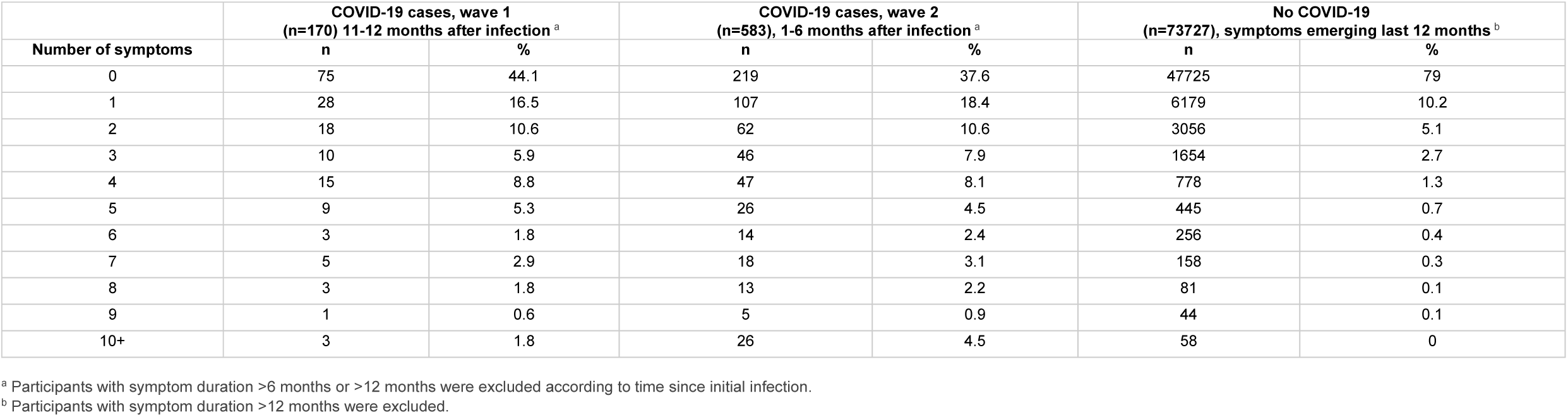
Number of symptoms reported in March 2021 among COVID-19 cases in wave 1, wave 2, and among subjects not receiving a COVID-19 diagnosis. ^a^

| Number of symptoms | COVID-19 cases, wave 1<br>(n=170) 11-12 months after infection <sup>a</sup> |  | COVID-19 cases, wave 2<br>(n=583), 1-6 months after infection <sup>a</sup> |  | No COVID-19<br>(n=73727), symptoms emerging last 12 months <sup>b</sup> |  |
| --- | --- | --- | --- | --- | --- | --- |
|  | n | % | n | % | n | % |
| 0 | 75 | 44.1 | 219 | 37.6 | 47725 | 79 |
| 1 | 28 | 16.5 | 107 | 18.4 | 6179 | 10.2 |
| 2 | 18 | 10.6 | 62 | 10.6 | 3056 | 5.1 |
| 3 | 10 | 5.9 | 46 | 7.9 | 1654 | 2.7 |
| 4 | 15 | 8.8 | 47 | 8.1 | 778 | 1.3 |
| 5 | 9 | 5.3 | 26 | 4.5 | 445 | 0.7 |
| 6 | 3 | 1.8 | 14 | 2.4 | 256 | 0.4 |
| 7 | 5 | 2.9 | 18 | 3.1 | 158 | 0.3 |
| 8 | 3 | 1.8 | 13 | 2.2 | 81 | 0.1 |
| 9 | 1 | 0.6 | 5 | 0.9 | 44 | 0.1 |
| 10+ | 3 | 1.8 | 26 | 4.5 | 58 | 0 |

<sup>a</sup> Participants with symptom duration >6 months or >12 months were excluded according to time since initial infection.
<sup>b</sup> Participants with symptom duration >12 months were excluded.

**Supplementary Table 7.** Bivariate tetrachoric correlation coefficients between 20 symptoms among all COVID-19 cases (n=774). Rare occurrences (myocarditis and kidney disease) were excluded.

|  | Fatigue | Fever | Shortness of breath | Cough | Reduced lung function | Joint pain | Muscle pain | Chest pain | Heart palpitation | Dizziness | Brain fog | Poor memory | Mood changes | Sleep problems | Headache | Depression | Anxiety | Skin rash | Hair loss | Altered smell or taste |
| --- | --- | --- | --- | --- | --- | --- | --- | --- | --- | --- | --- | --- | --- | --- | --- | --- | --- | --- | --- | --- |
| Fatigue | 1 | 0.17 | 0.64 | 0.33 | 0.37 | 0.42 | 0.41 | 0.49 | 0.65 | 0.59 | 0.7 | 0.73 | 0.59 | 0.52 | 0.61 | 0.51 | 0.49 | 0.38 | 0.56 | 0.33 |
| Fever | 0.17 | 1 | 0.09 | -0.36 | -0.38 | 0.36 | 0.38 | 0.36 | 0.23 | 0.39 | 0.34 | 0.23 | 0.22 | 0.05 | 0.42 | -0.33 | -0.36 | 0.27 | 0.22 | 0.25 |
| Shortness of breath | 0.64 | 0.09 | 1 | 0.47 | 0.58 | 0.46 | 0.48 | 0.56 | 0.45 | 0.38 | 0.45 | 0.57 | 0.32 | 0.38 | 0.54 | 0.24 | 0.3 | 0.35 | 0.32 | 0.25 |
| Cough | 0.33 | -0.36 | 0.47 | 1 | 0.38 | 0.02 | 0.22 | 0.18 | 0.18 | -0.01 | 0.12 | 0.19 | -0.04 | 0.26 | 0.23 | 0.07 | 0.12 | 0.17 | 0.27 | 0.15 |
| Reduced lung function | 0.37 | -0.38 | 0.58 | 0.38 | 1 | 0.12 | 0.03 | 0.4 | 0.2 | 0.26 | 0.31 | 0.37 | -0.05 | 0.05 | 0.23 | 0.1 | 0.27 | 0.09 | 0 | 0.13 |
| Joint pain | 0.42 | 0.36 | 0.46 | 0.02 | 0.12 | 1 | 0.67 | 0.38 | 0.51 | 0.4 | 0.41 | 0.4 | 0.37 | 0.36 | 0.44 | 0.24 | 0.33 | 0.36 | 0.18 | 0.21 |
| Muscle pain | 0.41 | 0.38 | 0.48 | 0.22 | 0.03 | 0.67 | 1 | 0.4 | 0.49 | 0.57 | 0.46 | 0.5 | 0.43 | 0.46 | 0.5 | 0.22 | 0.25 | 0.34 | 0.39 | 0.25 |
| Chest pain | 0.49 | 0.36 | 0.56 | 0.18 | 0.4 | 0.38 | 0.4 | 1 | 0.35 | 0.4 | 0.45 | 0.46 | 0.11 | 0.24 | 0.52 | -0.05 | 0.16 | 0.08 | 0.41 | 0.05 |
| Heart palpitation | 0.65 | 0.23 | 0.45 | 0.18 | 0.2 | 0.51 | 0.49 | 0.35 | 1 | 0.66 | 0.63 | 0.65 | 0.54 | 0.61 | 0.57 | 0.4 | 0.44 | 0.28 | 0.49 | 0.31 |
| Dizziness | 0.59 | 0.39 | 0.38 | -0.01 | 0.26 | 0.4 | 0.57 | 0.4 | 0.66 | 1 | 0.65 | 0.63 | 0.51 | 0.55 | 0.65 | 0.27 | 0.3 | 0.41 | 0.4 | 0.24 |
| Brain fog | 0.7 | 0.34 | 0.45 | 0.12 | 0.31 | 0.41 | 0.46 | 0.45 | 0.63 | 0.65 | 1 | 0.84 | 0.58 | 0.6 | 0.6 | 0.43 | 0.33 | 0.48 | 0.41 | 0.27 |
| Poor memory | 0.73 | 0.23 | 0.57 | 0.19 | 0.37 | 0.4 | 0.5 | 0.46 | 0.65 | 0.63 | 0.84 | 1 | 0.45 | 0.54 | 0.52 | 0.41 | 0.39 | 0.38 | 0.44 | 0.41 |
| Mood changes | 0.59 | 0.22 | 0.32 | -0.04 | -0.05 | 0.37 | 0.43 | 0.11 | 0.54 | 0.51 | 0.58 | 0.45 | 1 | 0.62 | 0.66 | 0.77 | 0.67 | 0.55 | 0.52 | 0.17 |
| Sleep problems | 0.52 | 0.05 | 0.38 | 0.26 | 0.05 | 0.36 | 0.46 | 0.24 | 0.61 | 0.55 | 0.6 | 0.54 | 0.62 | 1 | 0.51 | 0.56 | 0.45 | 0.15 | 0.46 | 0.2 |
| Headache | 0.61 | 0.42 | 0.54 | 0.23 | 0.23 | 0.44 | 0.5 | 0.52 | 0.57 | 0.65 | 0.6 | 0.52 | 0.66 | 0.51 | 1 | 0.32 | 0.3 | 0.51 | 0.3 | 0.28 |
| Depression | 0.51 | -0.33 | 0.24 | 0.07 | 0.1 | 0.24 | 0.22 | -0.05 | 0.4 | 0.27 | 0.43 | 0.41 | 0.77 | 0.56 | 0.32 | 1 | 0.88 | 0.4 | 0.28 | 0.02 |
| Anxiety | 0.49 | -0.36 | 0.3 | 0.12 | 0.27 | 0.33 | 0.25 | 0.16 | 0.44 | 0.3 | 0.33 | 0.39 | 0.67 | 0.45 | 0.3 | 0.88 | 1 | 0.39 | 0.46 | 0.07 |
| Skin rash | 0.38 | 0.27 | 0.35 | 0.17 | 0.09 | 0.36 | 0.34 | 0.08 | 0.28 | 0.41 | 0.48 | 0.38 | 0.55 | 0.15 | 0.51 | 0.4 | 0.39 | 1 | 0.21 | 0.33 |
| Hair loss | 0.56 | 0.22 | 0.32 | 0.27 | 0 | 0.18 | 0.39 | 0.41 | 0.49 | 0.4 | 0.41 | 0.44 | 0.52 | 0.46 | 0.3 | 0.28 | 0.46 | 0.21 | 1 | 0.2 |
| Altered smell or taste | 0.33 | 0.25 | 0.25 | 0.15 | 0.13 | 0.21 | 0.25 | 0.05 | 0.31 | 0.24 | 0.27 | 0.41 | 0.17 | 0.2 | 0.28 | 0.02 | 0.07 | 0.33 | 0.2 | 1 |

**Supplementary Figure 1.**
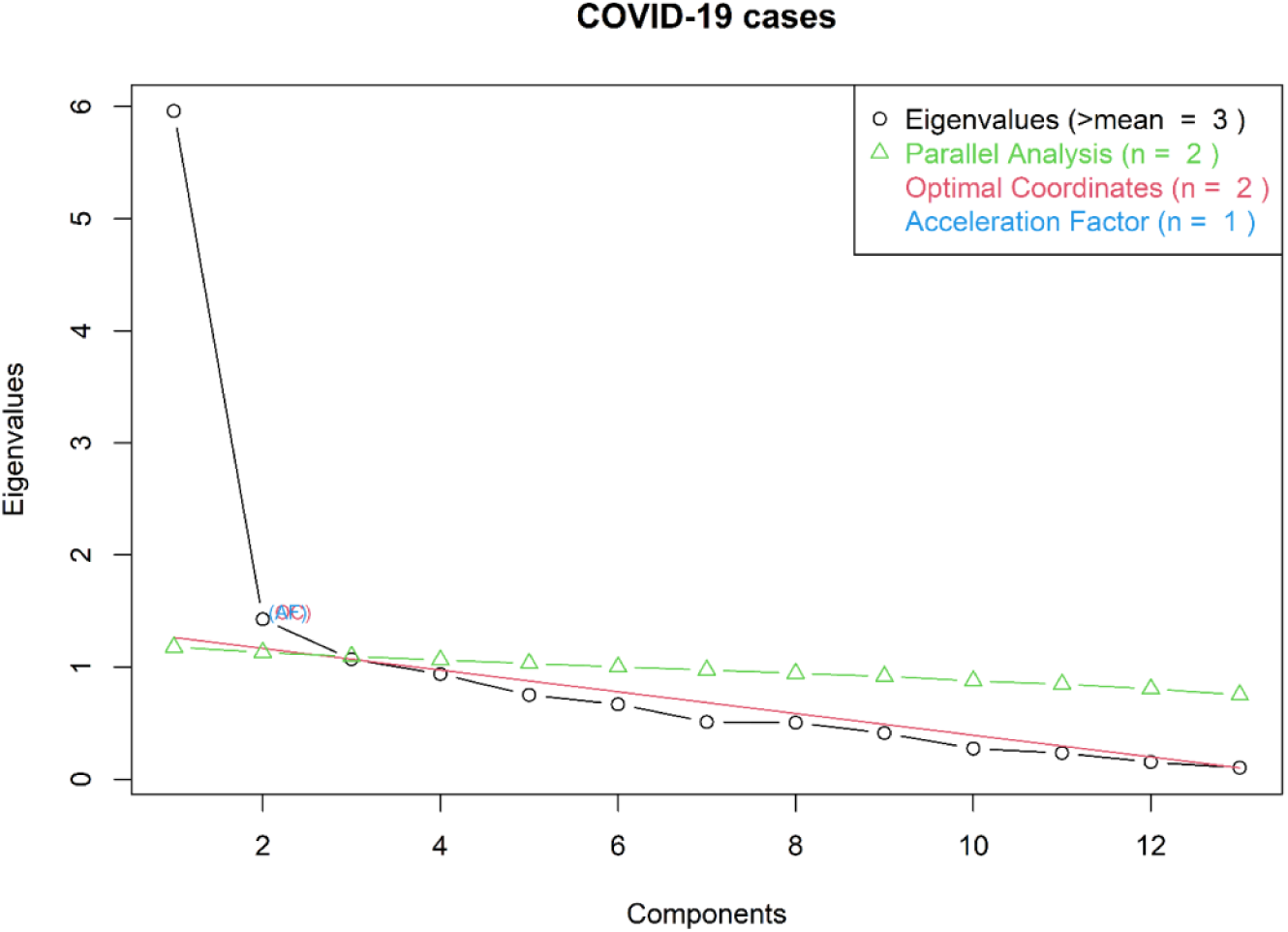
Scree plot from R package *nFactors*. The first three factors had eigenvalues eigenvalues 6.0, 1.4 and 1.1. The Kaiser criterion (eigenvalue >1) suggested three factors and Horn’s parallel analysis suggested that two factors should be retained in the model. Parallel analysis was run with 100 replications of the correlation matrix.

**Supplementary Figure 2.**
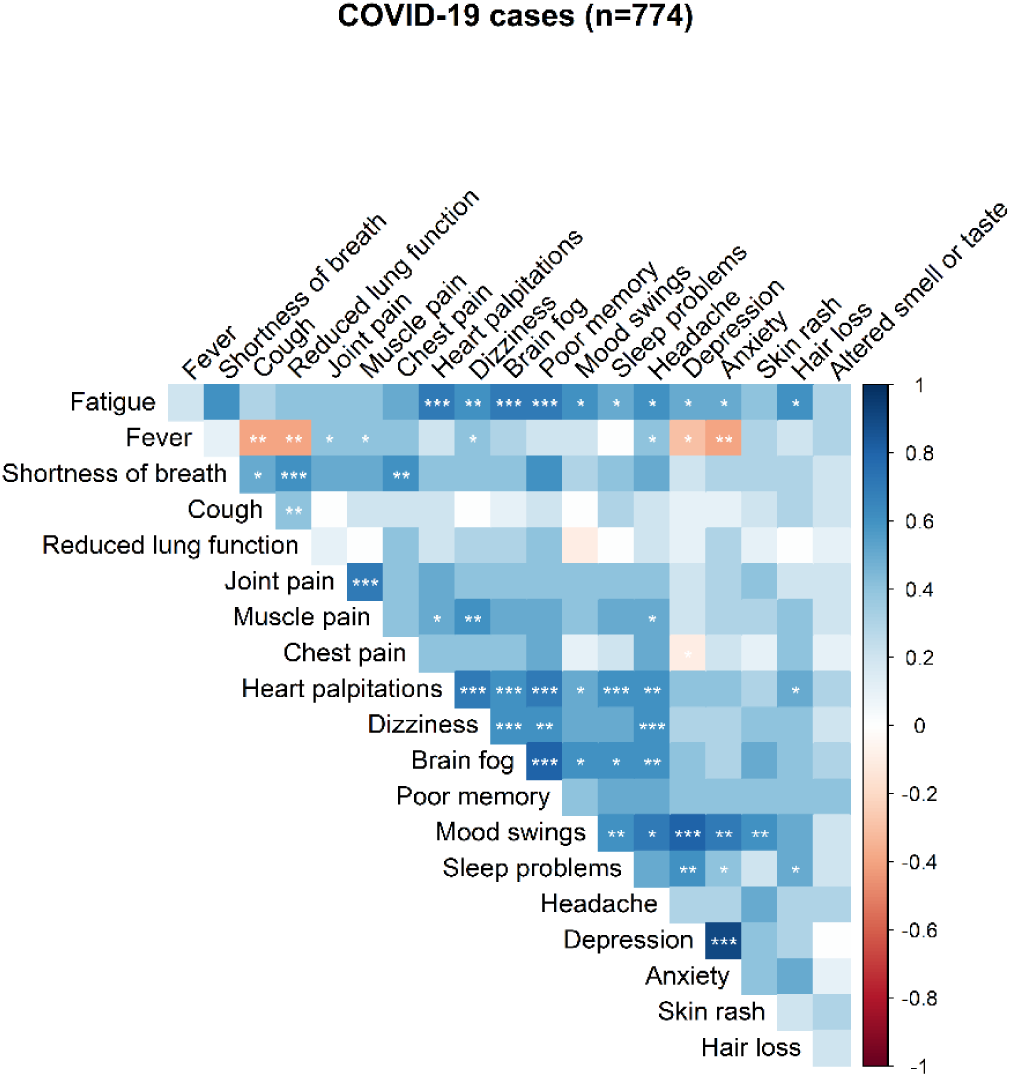
Bivariate tetrachoric correlations between symptoms reported in March 2021 among COVID-19 cases (n=774). Rare occurrences (myocarditis and kidney disease) were excluded. The strength of correlation coefficients is indicated by the colour panel (right). Intensity of red colours indicate increasing negative correlation coefficients, while intensity of blue colours indicates increasing positive correlation coefficients. Asterisks indicating significant correlations (*** for p<.001; ** for p<.01; * for p<.05).

## Notes

### Competing Interest Statement

The authors have declared no competing interest.

### Funding Statement

This work was funded by the Norwegian Research Council Centres of Excellence Funding Scheme (no. 262700) and by the Norwegian Institute of Public Health (NIPH).

### Author Declarations

This work was approved by The Regional Committee for Medical and Health Research Ethics, South East Norway C, no. 127708.

